# COVID-19 and its clinical severity are associated with alterations of plasma sphingolipids and enzyme activities of sphingomyelinase and ceramidase

**DOI:** 10.1101/2022.01.19.22269391

**Authors:** Christiane Mühle, Andreas Kremer, Marcel Vetter, Jonas Schmid, Susanne Achenbach, Fabian Schumacher, Bernd Lenz, Céline Cougoule, Nicolas Hoertel, Alexander Carpinteiro, Erich Gulbins, Burkhard Kleuser, Johannes Kornhuber

## Abstract

In the current pandemic caused by the severe acute respiratory syndrome coronavirus 2 (SARS-CoV-2; COVID-19), a better understanding of the underlying mechanisms is essential to reduce morbidity and mortality and treat post-COVID-19 disease. Here, we analyzed alterations of sphingolipids and their metabolizing enzymes in 125 men and 74 women tested positive for SARS-CoV-2 and hospitalized with mild, moderate or severe symptoms or after convalescence.

The activities of acid and neutral sphingomyelinases (ASM, NSM), which hydrolyze sphingomyelin to ceramide, were significantly increased in COVID-19 patients, while the activity of neutral ceramidase (NC), which hydrolyzes ceramide to sphingosine, was reduced. These alterations could each contribute to elevated ceramide levels in patients. Accordingly, liquid chromatography tandem-mass spectrometry (LC-MS/MS) yielded increased levels of ceramides 16:0 and 18:0 with highest levels in severely affected patients and similar effects for dihydroceramides 16:0 and 18:0, whereas levels of (dihydro-)ceramides 24:0 were reduced. Furthermore, sphingomyelin 20:0; 22:0 and 24:0 as substrates of ASM and NSM as well as their dihydrosphingomyelin counterparts were reduced in patients as well as sphingosine-1-phosphate further downstream of NC activity. Effects of NSM, NC, ceramides and sphingomyelins remained significant after Bonferroni correction. SARS-CoV-2 antibody levels in convalescent patients were associated with age but none of the sphingolipid parameters. Based on our data, COVID-19 is associated with a dysregulation of sphingolipid homeostasis in a severity-dependent manner, particularly focused around a reduction of sphingomyelins and an accumulation of ceramides by increased enzyme activities leading to ceramide elevation (ASM, NSM) combined with a decreased activity of enzymes (NC) reducing ceramide levels. The potential of a combined sphingolipid/enzyme pattern as a diagnostic and prognostic marker and therapeutic target deserves further exploration.

## Introduction

To contain the current pandemic caused by the severe acute respiratory syndrome coronavirus 2 (SARS-CoV-2), a better understanding of the underlying mechanisms and the high interindividual differences in susceptibility is essential. A number of recent studies point to changes in the sphingolipid metabolism (Kornhuber et al. 2021). Sphingolipids such as sphingomyelin (SM) and ceramide (Cer) not only serve as physical membrane components and as ligands on their own, but they also influence the local composition and fluidity of the plasma membrane and thereby the localization and activity of proteins involved in receptor-mediated signaling (Hannun and Obeid 2018). In patients with SARS-CoV-2 (COVID-19 patients), the Cer-signaling pathway was noted to be upregulated (Alexander et al. 2021). Cer concentrations were found to be increased in the plasma of COVID-19 patients per se (Marin-Corral et al. 2021) (Lee et al. 2021a) and an additional increase was seen in patients with severe respiratory symptoms (Khodadoust 2021; Torretta et al. 2021). Furthermore, reduced serum sphingosine levels were highly associated with symptomatic COVID-19 compared to asymptomatic SARS-CoV-2 antibody-positive donors (Janneh et al. 2021). Another metabolomics study found plasma metabolites that negatively correlated with diseases severity to be enriched for metabolism of sphingolipids and glycerophospholipids. This group also identified an emerging metabolically dominant natural killer cell subpopulation with increased metabolic activity in sphingolipid metabolism (Lee et al. 2021a).

Sphingolipid synthesis and conversions are regulated by a complex interconnected pathway of enzymes to keep homeostasis. Sphingomyelinases such as acid (ASM) or neutral (NSM) sphingomyelinase catalyze the hydrolysis of SM to Cer and phosphorylcholine under acidic or neutral pH conditions, respectively, and can thereby lead to an increase in Cer levels. In contrast, ceramidases such as acid (AC) and neutral (NC) ceramidase catalyze the degradation of Cer to sphingosine and fatty acid within their specific optimal pH range (Figure 1) (Hannun and Obeid 2018). Thus, these and further enzymes are crucial in controlling Cer levels. Many antidepressants (and other drugs) of different groups including selective serotonin reuptake inhibitors (SSRIs) have been found to indirectly inhibit ASM (Kornhuber et al. 2011) by accumulation within the lysosome and detachment of ASM from the inner lysosomal membrane exposing the enzyme to proteolytic degradation (Hurwitz et al. 1994). These drugs were therefore named functional inhibitors of ASM (FIASMA, (Kornhuber et al. 2010)) and include commonly used antidepressants like fluoxetine, amitriptyline and fluvoxamine.

**Figure 1:**
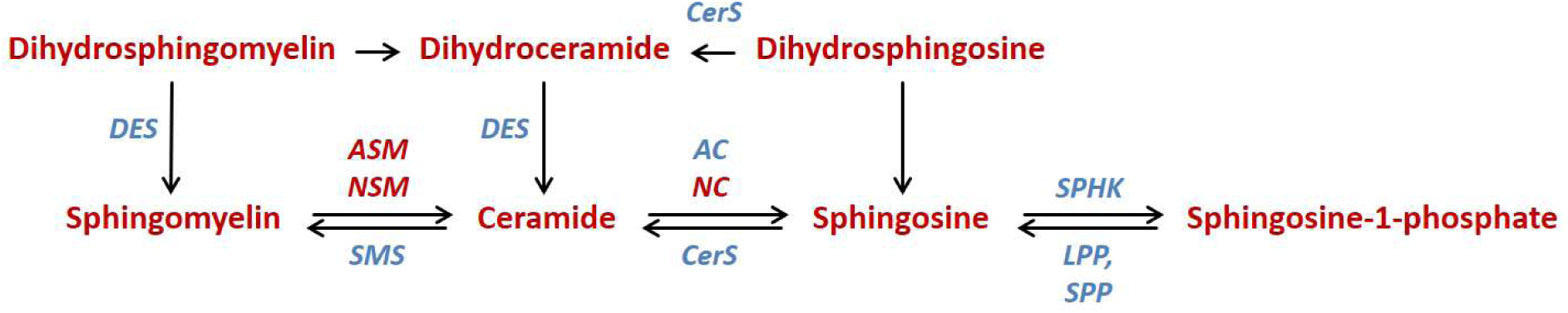
Biochemical pathway of sphingolipids with central ceramide (analyzed sphingolipid species in red, decreasing abundance from left to right) and enzymes catalyzing conversions (italicized, analyzed species in red). AC, NC: Acid and neutral ceramidases; ASM, NSM: Acid and neutral sphingomyelinases; CerS: Ceramide synthase, DES: Dihydroceramide desaturase; LPP: Lipid phosphate phosphatase; SMS: Sphingomyelin synthase; SPHK: Sphingosine kinase; SPP: Sphingosine phosphate phosphatase.

Interestingly, an antiviral effect of FIASMAs against SARS-CoV, Middle East Respiratory Syndrome (MERS)-CoV and also especially SARS-CoV-2 has been demonstrated by various approaches *in silico, in vitro* and *in vivo* as summarized in recent reviews (Kornhuber et al. 2021; Le Corre and Loas 2021) (Loas and Le Corre 2021).

SARS-CoV-2 is thought to activate the ASM/Cer system, resulting in the formation of Cer-enriched membrane domains (Carpinteiro et al. 2020) (Schneider-Schaulies et al. 2021) and clustering of angiotensin-converting enzyme 2 (ACE2), the cellular receptor of SARS-CoV-2. This in turn facilitates viral entry and infection and consequently the release of proinflammatory cytokines (Tornquist et al. 2021). Cell biological studies indicate a reduced rate of infection of cell lines or primary nasal epithelial cells by SARS-CoV-2 after administration of FIASMAs fluoxetine or ambroxol and restored infection after reconstitution of Cer in these cells (Carpinteiro et al. 2020; Carpinteiro et al. 2021) (Schloer et al. 2020).

From the treatment perspective, a multicenter, retrospective, observational study on hospitalized patients with COVID-19 has indicated a reduced risk of intubation or death in patients taking antidepressants, particularly fluoxetine (Hoertel et al. 2021b). Similarly, a large retrospective cohort study observed reduced mortality in patients treated with fluoxetine or fluvoxamine compared to matched patients not treated with these or any other SSRIs (Oskotsky et al. 2021). Two further randomized placebo-controlled trials and a prospective real-world study support the observation of a lower risk of clinical deterioration over two weeks of treatment with fluvoxamine (Lenze et al. 2020; Reis et al. 2022; Seftel and Boulware 2021). Finally, an open-label prospective cohort trial with matched controls reported that fluvoxamine treatment in addition to the standard therapy was significantly and substantially associated with reduced mortality (Calusic et al. 2021). Importantly, the beneficial clinical effects in COVID-19 are predominantly mediated by FIASMAs (Hoertel et al. 2021a; Hoertel et al. 2021c; Kornhuber et al. 2021).

Although several studies have reported alterations of sphingolipid patterns in COVID-19 patients so far, sphingolipid metabolizing enzymes have been mostly neglected as potential biomarkers. In addition to sphingolipidomics, we therefore aimed at analyzing the activities of ASM but also NSM and NC which are detectable in human plasma and serum with the aim of developing biomarkers to predict and track disease progression and add to the understanding of underlying mechanisms to improve therapeutic approaches. Moreover, our study includes convalescent individuals recovered from a COVID-19 infection and is based on sex-balanced groups with explorative sex-specific analysis.

## Materials and methods

### Study description

Patients with COVID-19 infection and convalescent patients (plasma donors) were recruited at the Universitätsklinikum Erlangen, Germany either during hospitalization (patients) or as part of a project to obtain convalescent plasma from individuals who have recovered from COVID-19 with the aim to treat infected patients with human antibodies. Diagnosis for COVID-19 was based on a positive PCR test for SARS-CoV-2; convalescent plasma donors had negative PCR results at the time of plasma donation. Patients were classified based on purely clinical parameters: Those with “mild” symptoms were hospitalized in the normal ward and did not require oxygen at later stages in contrast to those with “moderate” and “severe” symptoms. Severely affected patients had a stay at the intensive care unit, some with extracorporeal membrane oxygenation, and/or died during the course of disease. The study was approved by the Ethics Committee of the Medical Faculty of the Friedrich-Alexander University Erlangen-Nürnberg (174_20B, April 30, 2020 for patients and 357_19B, October 18^th^, 2019 for convalescent plasma donors). All participants provided written informed consent.

Blood samples were collected in most cases in the morning in fastened state as part of routine blood work at initial diagnosis and during clinical follow-up. Plasma citrate vials were centrifuged (10 min and 2000 g at room temperature), and plasma was aliquoted and placed into storage at –80°C for later enzyme activity assays and liquid chromatography tandem-mass spectrometry (LC-MS/MS). Leukocyte and thrombocyte counts as well as C-reactive protein, were quantified at the Central Laboratory of the Universitätsklinikum Erlangen, Germany (DIN EN ISO 15189 accredited) from separately collected serum and EDTA vials.

### Enzyme activity assays for sphingomyelinase and ceramidase

The activity of sphingolipid enzymes was quantified using fluorescent substrates, BODIPY-FL-C_12_-Sphingomyelin (D-7711, Invitrogen, Carlsbad, CA, USA/Life Technologies, Grand 15 Island, NY, USA) for S-ASM and NSM (Kalinichenko et al. 2021a; Kalinichenko et al. 2021b) and NBD-C_12_-ceramide (Cay10007958-1, Cayman Chemical, via Biomol GmbH, Hamburg, Germany) for neutral ceramidase (Kalinichenko et al. 2021c) as described previously (Mühle and Kornhuber 2017). Briefly, the reaction was performed in 96 well polystyrene plates with 58 or 50 pmol fluorescently labelled sphingomyelin or Cer, respectively, in a buffer mix totaling 50 μl in volume. The reaction was initiated by the addition of 6 μl of a 1:10 dilution of plasma in physiological 154 mM NaCl solution. After incubation at 37°C for 6-48 hours depending on the enzyme, reactions were stopped by freezing at –20°C and stored until further processing. For direct chromatography, 1.5 μl of the reaction was spotted directly without further purification on silica gel 60 thin layer chromatography plates (ALUGRAM SIL G, 818232, Macherey-Nagel, Düren, Germany). Product and uncleaved substrate were separated using ethyl acetate with 1% (v/v) acetic acid as a solvent for all enzymes. Spot intensities were detected on a Typhoon Trio scanner and quantified using the ImageQuant software (GE Healthcare Life Sciences, Buckinghamshire, UK). All enzyme activity assays were carried out with four replicate dilutions of each sample and using the same lot of reagents and consumables and performed by a single operator.

### Sphingolipid quantification by liquid chromatography tandem-mass spectrometry (LC-MS/MS)

MS-grade methanol was added to plasma samples (500 μl per 20 μl) before shipment of samples on dry ice. Sphingolipids were extracted as described previously (Gulbins et al. 2018). To this end 1 mL methanol/chloroform (1:1, v:v) containing the internal standards d_7_-dihydrosphingosine (d_7_-dHSph), d_7_-sphingosine (d_7_-Sph), d_7_-sphingosine 1-phosphate (d_7_-S1P), 17:0 Cer (d18:1/17:0) and d_31_-16:0 sphingomyelin (d18:1/16:0-d_31_) (all Avanti Polar Lipids, Alabaster, AL, USA) were added. Extraction was facilitated by incubation at 48°C with gentle shaking (120 rpm) overnight. To reduce interference from plasma glycerolipids, samples were saponified with 150 μL 1 M methanolic KOH for 2 h at 37°C with gentle shaking (120 rpm) followed by neutralization with 12 μL glacial acetic acid. After centrifugation at 2,200 g for 10 min at 4°C, organic supernatants were evaporated to dryness using a Savant SpeedVac concentrator (Thermo Fisher Scientific, Dreieich, Germany). Dried residues were reconstituted in 200 μL acetonitrile/methanol/water (47.5:47.5:5 (v:v:v), 0.1% formic acid) and subjected to LC-MS/MS sphingolipid quantification applying the multiple reaction monitoring (MRM) approach. Chromatographic separation was achieved on a 1290 Infinity II HPLC (Agilent Technologies, Waldbronn, Germany) equipped with a Poroshell 120 EC-C8 column (3.0 × 150 mm, 2.7 μm; Agilent Technologies) guarded by a pre-column (3.0 × 5 mm, 2.7 μm) of identical material. MS/MS analyses were carried out using a 6495 triple-quadrupole mass spectrometer (Agilent Technologies) operating in the positive electrospray ionization mode (ESI+). Chromatographic conditions and settings of the ESI source and MS/MS detector have been published elsewhere (Naser et al. 2020).

The following mass transitions were recorded (qualifier product ions in parentheses): <u>long-chain bases (LCB):</u> *m/z* 300.3 → 282.3 (252.3) for Sph, *m/z* 302.3 → 284.3 (254.3) for dHSph, *m/z* 307.3 → 289.3 (259.3) for d_7_-Sph, *m/z* 309.4 → 291.3 (261.3) for d_7_-dHSph, *m/z* 380.3 → 264.3 (82.1) for S1P, and *m/z* 387.3 → 271.3 (82.1) for d_7_-S1P; <u>(Cer):</u> *m/z* 520.5 → 264.3 (282.3) for 16:0 Cer, *m/z* 534.5 → 264.3 (282.3) for 17:0 Cer, *m/z* 548.5 → 264.3 (282.3) for 18:0 Cer, *m/z* 576.6 → 264.3 (282.3) for 20:0 Cer, *m/z* 604.6 → 264.3 (282.3) for 22:0 Cer, *m/z* 630.6 → 264.3 (282.3) for 24:1 Cer, and *m/z* 632.6 → 264.3 (282.3) for 24:0 Cer; <u>dihydroceramides (dHCer):</u> *m/z* 540.5 → 522.6 (284.3) for 16:0 dHCer, *m/z* 568.5 → 550.5 (284.3) for 18:0 dHCer, *m/z* 596.6 → 578.6 (284.3) for 20:0 dHCer, *m/z* 624.6 → 606.6 (284.3) for 22:0 dHCer, *m/z* 650.7 → 632.7 (284.3) for 24:1 dHCer, and *m/z* 652.7 → 634.6 (284.3) for 24:0 dHCer; <u>sphingomyelins (SM):</u> *m/z* 703.6 → 184.1 (86.1) for 16:0 SM, *m/z* 731.6 → 184.1 (86.1) for 18:0 SM, *m/z* 734.6→ 184.1 (86.1) for 16:0-d_31_ SM, *m/z* 759.6 → 184.1 (86.1) for 20:0 SM, *m/z* 787.7 → 184.1 (86.1) for 22:0 SM, *m/z* 813.7 → 184.1 (86.1) for 24:1 SM, and *m/z* 815.7 → 184.1 (86.1) for 24:0 SM; <u>dihydrosphingomyelins (dHSM):</u> *m/z* 705.6 → 184.1 (86.1) for 16:0 dHSM, *m/z* 733.6 → 184.1 (86.1) for 18:0 dHSM, *m/z* 761.6 → 184.1 (86.1) for 20:0 dHSM, *m/z* 789.7→ 184.1 (86.1) for 22:0 dHSM, *m/z* 815.7 → 184.1 (86.1) for 24:1 dHSM, and *m/z* 817.7 → 184.1 (86.1) for 24:0 dHSM.

Peak areas of Cer, dHCer, SM and dHSM subspecies were normalized to those of their internal standards (17:0 Cer for Cer and dHCer subspecies; 16:0-d_31_ SM for SM and dHSM subspecies) followed by external calibration in the range of 1 fmol to 5 pmol (dHCer) or 50 pmol (Cer, SM, dHSM) on column. dHSph, Sph and S1P were directly quantified via their deuterated internal standards d_7_-dHSph (0.125 pmol on column), d_7_-Sph (0.25 pmol on column) and d_7_-S1P (0.125 pmol on column). Quantification was performed with MassHunter Software (Agilent Technologies).

### Determination of SARS-CoV-2 antibody levels

Levels of antibodies against SARS-CoV-2 in plasma donors (patients after convalescence) were determined by two enzyme immunoassays: LIAISON® SARS-CoV-2 S1/S2 IgG (DiaSorin Deutschland GmbH, Dietzenbach, Germany, n=27), a semiquantitative assay, and SARS-CoV-2-EIA (EUROIMMUN AG, Lübeck, Germany, n=23). Antibody levels were converted to t-scores for each test to be able to use the combined values as one variable.

### Statistical analyses

For statistics, SPSS for Windows 28.0 (SPSS Inc., Chicago, IL) was used and means with standard deviation (SD) are reported (SPSS custom tables function) where the t-test was applied to analyze differences. Graphs were prepared using GraphPad Prism 8.4.3 (Graph Pad Soft-ware Inc., San Diego, CA, USA). In the case of missing data points, study subjects were excluded from the specific analyses. To determine the impact of enzyme activities and sphingolipids on disease status and severity (0 = convalescent, 1 = mild, 2 = moderate, 3 = severe symptoms), stepwise linear regression models were used with sex and age as predictors in the first step and the sphingolipid parameter in the second step yielding a standardized B coefficient, p for the change in F and the change in R^2^ (see Table 1). The results were validated using bias-corrected and accelerated bootstrap (1,000 resamples). For visualization, further confirmation and determination of odds ratios, multinomial logistic regression was performed with sex and age as cofactors and z-scores of raw values for the factors based on mean and SD of the convalescent group (see Figure 1 and Table S2). Nominal p values are provided in the text and tables and p < 0.05 for two-tailed tests was considered significant. Bonferroni correction was applied for multiple testing (in total for 3 enzymes and 31 sphingolipid species). Because of the importance of sex differences in science generally (Tannenbaum et al. 2019) and sex differences for some parameters as well as known sex-dependent effects of sphingolipid enzymes (Mühle et al. 2014; Muhle et al. 2019b; Mühle et al. 2018), we also analyzed men and women separately.

**Table 1:** COVID-19 severity prediction by sphingolipids and sphingolipid metabolism enzymes in combination with sex and age as predictors (linear regression model coding 0 = convalecent, 1 = mild, 2 = moderate, 3 = severe symptoms). See table S5 for sex-separated analysis.

| Predictor | n | change | standardized B | p for change in F | change in R<sup>2</sup> |
| --- | --- | --- | --- | --- | --- |
| Plasma_ASM | 199 | ↑ | 0.136 | 0.024 | 0.018 |
| **Plasma_NSM** | 199 | ↑ | 0.379 | **6.25 × 10<sup>-11</sup>** | 0.135 |
| **Plasma_NC** | 199 | ↓ | -0.225 | **2.15 × 10<sup>-4</sup>** | 0.047 |
| dHSM_16:0 | 198 | ↓ | -0.134 | 0.024 | 0.018 |
| dHSM_18:0 | 198 |  | 0.069 | 0.244 | 0.005 |
| **dHSM_20:0** | 198 | ↓ | -0.276 | **5.02 × 10<sup>-6</sup>** | 0.070 |
| **dHSM_22:0** | 198 | ↓ | -0.270 | **1.23 × 10<sup>-5</sup>** | 0.064 |
| **dHSM_24:0** | 100 | ↓ | -0.536 | **4.85 × 10<sup>-9</sup>** | 0.225 |
| dHSM_24:1 | 198 |  | -0.087 | 0.149 | 0.007 |
| dHSM_total | 198 | ↓ | -0.150 | 0.012 | 0.022 |
| SM_16:0 | 198 | ↓ | -0.178 | 0.003 | 0.032 |
| SM_18:0 | 198 |  | -0.073 | 0.220 | 0.005 |
| **SM_20:0** | 198 | ↓ | -0.408 | **6.41 × 10<sup>-12</sup>** | 0.148 |
| **SM_22:0** | 198 | ↓ | -0.351 | **1.22 × 10<sup>-8</sup>** | 0.106 |
| **SM_24:0** | 198 | ↓ | -0.323 | **1.31 × 10<sup>-7</sup>** | 0.092 |
| SM_24:1 | 198 |  | -0.091 | 0.127 | 0.008 |
| **SM_total** | 198 | ↓ | -0.255 | **1.80 × 10<sup>-5</sup>** | 0.062 |
| **dHCer_16:0** | 198 | ↑ | 0.193 | **0.001** | 0.037 |
| **dHCer_18:0** | 198 | ↑ | 0.225 | **1.51 × 10<sup>-4</sup>** | 0.049 |
| dHCer_20:0 | 198 |  | 0.067 | 0.257 | 0.005 |
| dHCer_22:0 | 198 |  | -0.073 | 0.220 | 0.005 |
| **dHCer_24:0** | 198 | ↓ | -0.254 | **2.85 × 10<sup>-5</sup>** | 0.059 |
| dHCer_24:1 | 198 |  | 0.044 | 0.459 | 0.002 |
| dHCer_total | 198 |  | -0.014 | 0.810 | 0.000 |
| **Cer_16:0** | 198 | ↑ | 0.279 | **2.45 × 10<sup>-6</sup>** | 0.074 |
| **Cer_18:0** | 198 | ↑ | 0.285 | **1.32 × 10<sup>-6</sup>** | 0.078 |
| Cer_20:0 | 198 | ↑ | 0.185 | 0.002 | 0.033 |
| Cer_22:0 | 198 | ↑ | 0.121 | 0.044 | 0.014 |
| **Cer_24:0** | 198 | ↓ | -0.190 | **0.001** | 0.035 |
| Cer_24:1 | 198 | ↑ | 0.187 | 0.002 | 0.032 |
| Cer_total | 198 |  | 0.083 | 0.168 | 0.007 |
| LCB_dHSph | 198 | ↓ | -0.180 | 0.003 | 0.031 |
| LCB_Sph | 198 | ↓ | -0.122 | 0.040 | 0.015 |
| LCB_S1P | 198 | ↓ | -0.139 | 0.023 | 0.018 |

ASM acid sphingomyelinase, NC neutral ceramidase, NSM neutral sphingomyelinase, LCB long chain base, dH dihydro, Sph sphingosine, S1P sphingosine-1-phosphate, Cer ceramide, SM sphingomyelin; n total sample size; for p < 0.05: ↑ increased and ↓ decreased in severely affected patients versus convalescent patients, all these remained p < 0.05 except for LCB-dHSph in Bootstrap analysis; predictor and p value in bold for those remaining significant after Bonferroni correction for multiple hypothesis testing.

## Results

### Demographic Characteristics

Patient blood was obtained during the intensive pandemic situation in the clinical emergency setting without specific selection or matching. We analyzed samples from 48 patients with mild symptoms (23 men; age: median 53, interquartile range 42-67 years), 50 patients with moderate symptoms (34 men; age: 67, 52-80 years) and 50 patients with severe symptoms (33 men; age: 69, 63-74 years) and compared these groups to 51 convalescent patients (35 men; age: 36, 29-53). As expected, the groups differed significantly in age (p < 0.001). Higher age is an established risk factor for hospitalization and severity of the COVID-19 infection. There was no significant difference in sex distribution between the groups (p = 0.107).

### Plasma sphingomyelinase and ceramidase activities are associated with disease severity

In the first step, we analyzed the relationship of plasma enzyme activities with COVID-19 severity (patients with severe > moderate > mild symptoms > convalescent). Linear regression analysis with age and sex as cofactors showed a significant association of disease severity with the activity of ASM (B = 0.136, p = 0.024) and NSM (B = 0.379, p = 6×10^−11^), respectively, both catalyzing the generation of Cer. Low activities of ASM and NSM were found in convalescent patients and higher activities in patients, with the highest NSM activities in severely affected patients (Table 1, S1). In contrast, the activity of NC hydrolyzing Cer was significantly decreased in COVID-19 patients in a severity-dependent manner (B = -0.225, p = 2×10^−4^, Table 1). The effects of NSM and NC remained significant after Bonferroni correction for multiple testing. In the linear regression analysis, age (all p < 0.005) but not sex (all p > 0.1) was significantly associated with the sphingolipid parameter in all models.

In a second analysis using multinomial regression, we were able to confirm and visualize that enzyme activities predict the symptom severity groups vs. the convalescent group and also severe vs. mild for NSM and NC and severe vs. moderate for NSM (Figure 2, Table S2). In this model, all three enzymes, ASM, NSM and NC, remained significant after Bonferroni correction.

**Figure 2:**
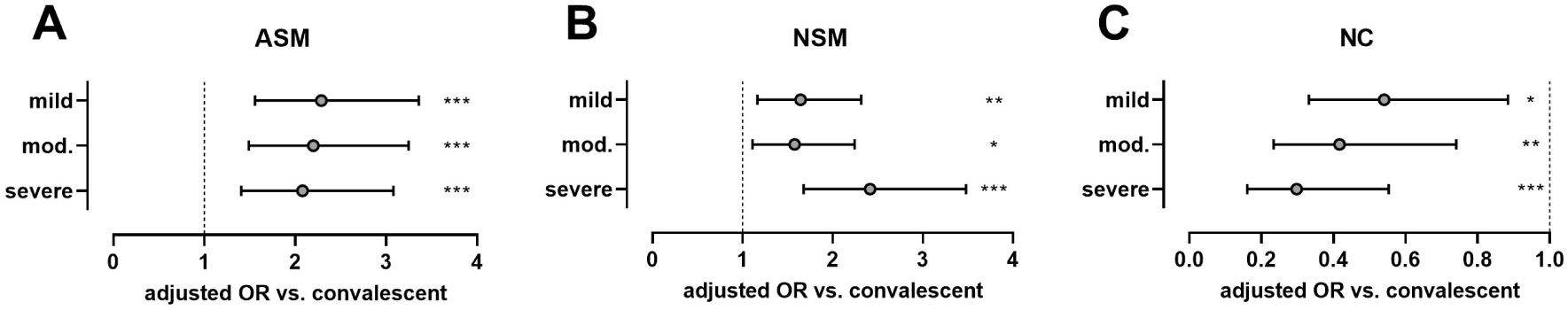
Odds ratios and significance of comparisons of patients with mild, moderate and severe symptoms with convalescent subjects (odds ratios with 95% confidence intervals as error bars) for sphingolipid metabolism enzymes from multinomial regression analysis of z-scores (calculated from raw values based on mean and standard deviation of the convalescent group) taking into account sex and age (see Table S2 for statistical information including further pair-wise comparisons and Figure S3 for visualization). *p < 0.05, **p < 0.01, ***p < 0.001

Although there were no sex differences in enzyme activities in the total cohort, we analyzed the effects for male and female participants separately and have observed very similar alterations in patients of both sexes (Table S5). However, the increase in ASM activity and decrease in NC activity in female were much stronger than in male patients.

### Plasma sphingolipid levels are associated with disease severity

In accordance with raised activity of ASM and NSM enzymes using sphingomyelin as a substrate, liquid chromatography tandem mass spectrometry of plasma samples revealed decreased levels of total plasma sphingomyelin (B = -0.255, p = 2×10^−5^) and of sphingomyelin species 20:0 (B = -0.408, p = 6×10^− 12^), 22:0 (B = -0.351, p = 1×10^−8^) and 24:0 (B = -0.323, p=1×10^−7^) as well as of dihydrosphingomyelin 20:0 (B = -0.276, p = 5×10^−6^), 22:0 (B = -0.270, p = 1×10^−5^) and 24:0 (B = -0.536, p = 5×10^−9^) in more severely affected COVID-19 patients. Accordingly, levels of Cers 16:0 (B = 0.279, p = 2×10^−6^) and 18:0 (B = 0.285, p = 1×10^−6^) as products of ASM and NSM were increased with highest levels in severely affected patients (Tables 1, S1). Moreover, levels of dihydroceramides 16:0 (B = 0.193, p = 0.001) and 18:0 (B = 0.225, p = 2×10^−4^) were also elevated in more severely affected COVID-19 patients. Only dihydroceramide (B = -0.254, p = 3×10^−5^) and Cer (B = -0.190, p = 0.001) with the specific fatty acid chain length 24:0 were decreased in patients with more severe infection symptoms. All of these 13 predictors remained significant after conservative Bonferroni correction for multiple hypothesis testing (bold in Table 1). Further factors with nominal significance in the statistical model were altered in the same manner, i.e. increased Cers (20:0, 22:0, 24:1) and decreased sphingomyelins (16:0) or dihydrosphingomyelins (16:0, total) in patients with more severe infection. Furthermore, dihydrosphingosine, sphingosine and sphingosine-1-phosphate were lower in more severely affected patients (Table 1).

Using multinomial regression, the same sphingolipid species remained significant to predict symptom severity group. In this model, sphingosine-1-phosphate was additionally significantly altered even after correction for multiple testing (Figure 3, Table S2).

**Figure 3:**
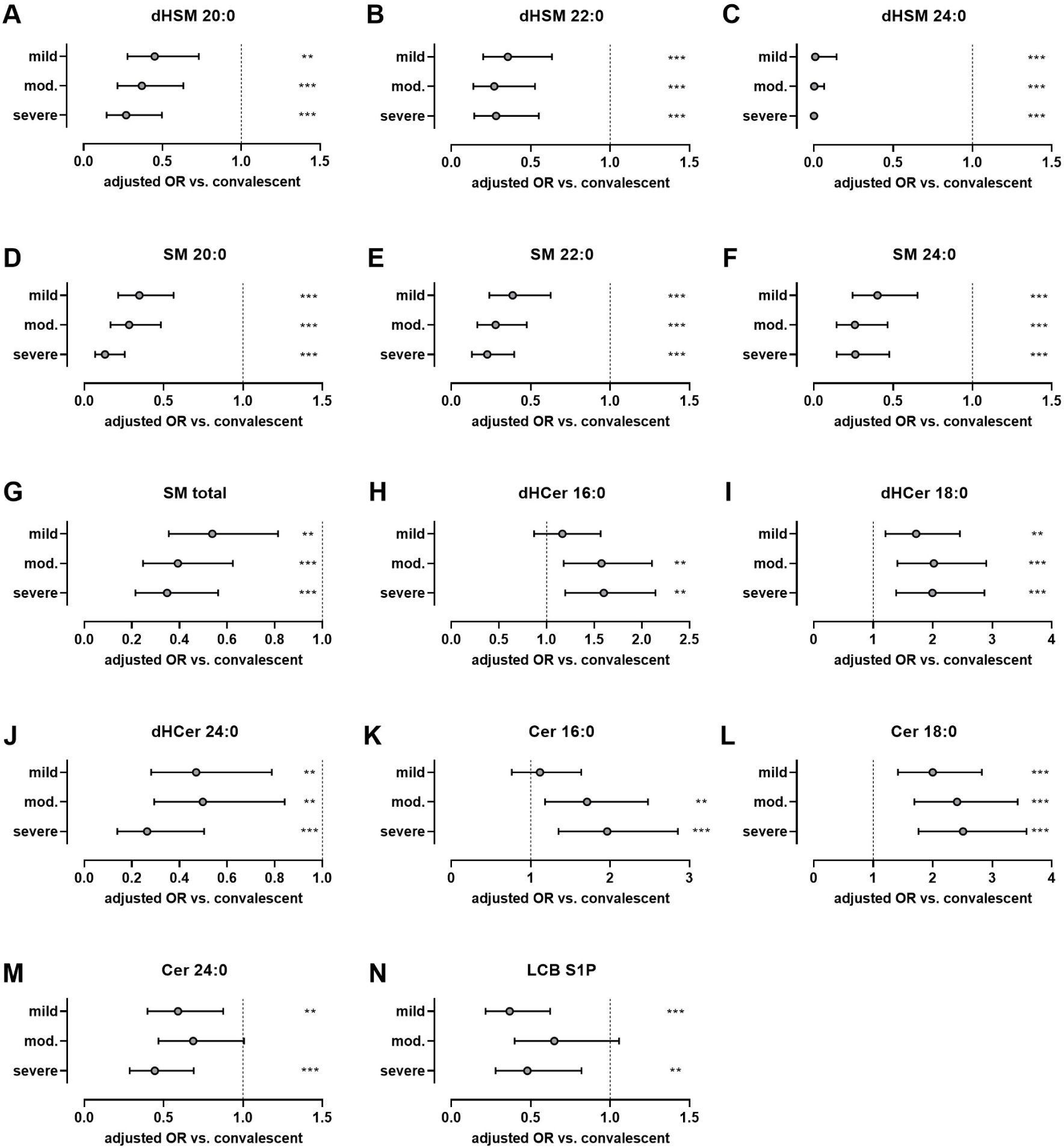
Odds ratios and significance of comparisons of patients with mild, moderate and severe symptoms with convalescent subjects (odds ratios with 95% confidence intervals as error bars) for sphingolipids (LC-MS/MS data, see Figure 1) from multinomial regression analysis of z-scores (calculated from raw values based on mean and standard deviation of the convalescent group) taking into account sex and age (see Table S2 for statistical information including further pair-wise comparisons and Figure S4 for visualization). **p < 0.01, ***p < 0.001

Although sphingolipid parameters did not differ significantly between male and female patients, sex-separated analysis revealed some differences in the strength (but not direction) of the effects. Some effects did not reach significance potentially due to the lower sample number. However, in females only, sphingomyelin 24:1 was reduced with increased symptom severity additionally to the other sphingomyelin species as a possible result of increased ASM and NSM activities. Contrariwise, the increases in Cer 20:0, 22:0 and 24:1 and decreases in dihydrospingosine and sphingosine were less pronounced or even absent in female compared to male patients (Table S5).

### Antibody levels in convalescent individuals

SARS-CoV-2 antibody levels of the 50 patients recovered from COVID-19 were highly age dependent (r_Spearman_ = 0.414, p = 0.003) and not associated with sphingolipid enzyme activities. In a linear regression model with age and sex as cofactors, none of the enzymes or sphingolipid parameters was associated with SARS-CoV-2 antibody levels (all p > 0.1).

## Discussion

Despite established risk factors of severe COVID-19 courses or mortality such as advanced age, obesity and pre-existing disease conditions, our understanding of biological mechanisms underlying the disease is very limited. However, such knowledge is needed for the developing of novel treatments with appropriate therapeutic response and low burden by adverse effects. So far, effective therapies are limited in their availability and the high costs (Kornhuber et al. 2021). Sphingolipids and their metabolizing enzymes are potential candidates for diagnostic and predictive biomarkers as well as therapeutic targets. To our knowledge, this is the first study to systematically assess these parameters including plasma ASM, NSM and NC activities in a larger sample of COVID-19 patients with different severities and to compare those to convalescent individuals.

Our data fit well with previous studies showing in COVID-19 patients an up-regulation of the Cer-signaling pathway (Alexander et al. 2021) and increased levels of Cer (Marin-Corral et al. 2021) (Abusukhun et al. 2021; Khodadoust 2021; Torretta et al. 2021) with Cer 24:0 as the solely decreased species (Torretta et al. 2021) as in our study. They are also in accordance with literature on increased dihydroceramide and decreased sphingomyelin in a cohort of 83 subjects (Torretta et al. 2021). We additionally show a decrease in dihydrosphingomyelin counterparts of sphingomyelin species. Although not remaining significant after conservative correction for multiple hypothesis testing, we have also observed reduced sphingosine (and additionally dihydrosphingosine) levels in patients, which agrees with the report of higher levels in symptomatic versus asymptomatic individuals (Janneh et al. 2021).

Despite the central role of enzymes involved in the regulation of Cer levels and the availability of inhibitors such as FIASMAs and thus their potential as prospective therapeutic targets, reports on these enzymes and particularly their activities are largely lacking so far apart from two studies at protein level and a very recent report on ASM activity: Our data on increased ASM activity in a larger cohort confirms the previously reported higher levels of ASM protein determined by ELISA in severely affected subjects from a total of 59 patients (Torretta et al. 2021) and the most recently observed increased serum ASM activities in 23 intensive care patients with COVID-19 assayed by a different method using mass spectrometry (Abusukhun et al. 2021). Whereas – to our knowledge – there are no data on NSM and NC activities in COVID-19 patients yet, our findings are also in perfect agreement with their contribution to increased ceramide and reduced sphingomyelin levels. Interestingly, Janneh et al. were able to quantify the enzyme AC in serum samples of symptomatic and asymptomatic individuals with COVID-19 infection versus healthy donors by western blot and observed clearly elevated and thus protective levels of AC in symptomatic patients (Janneh et al. 2021) in consent with increased Cer and with our increased NC activities. However, we were not able to determine any detectable AC activity in human serum or plasma samples so far compared to readily quantifiable activities in various mouse tissues.

Altogether, the altered activities in sphingolipid metabolizing enzymes observed here, i.e. increased activities of ASM and NSM catalyzing the production of Cer from sphingomyelin combined with decreased activities AC and NC catalyzing the degradation of Cer seem to be causative to the changes in plasma sphingolipid: decreased concentrations of (dihydro-)sphingomyelin species, increased concentrations of (dihydro-)Cers and decreased concentrations of (dihydro-)sphingosine and sphingosine-1-phosphate. These alterations in turn have been discussed to associate with many of the clinical phenomena observed in COVID-19 (Kornhuber et al. 2021).

Viral entry of SARS-CoV-2 is mediated via binding of the spike glycoprotein to its cellular receptor angiotensin-converting enzyme 2 (ACE2) within the host cell membrane (Wang et al. 2020). The highly hydrophobic Cer molecules associate spontaneously with gel-like Cer-enriched membrane domains which allow clustering, aggregation and reorganization of receptors (Kolesnick et al. 2000). Trapping and clustering of ACE2 in these platforms could amplify signaling and facilitate internalization of ACE2 and SARS-CoV-2 into endosomes (Carpinteiro et al. 2021) in addition to other potential mechanism increasing infection through the ASM/Cer system (Kornhuber et al. 2021).

Interestingly, the reduced levels of dihydrosphingomyelin in our more severely affected COVID-19 patients could also contribute to membrane changes favoring viral entrance, since dihydrosphingomyelin was reported to impair HIV-1 infection by rigidifying liquid-ordered membrane domains (Vieira et al. 2010). This minor component of lipid rafts in mammalian cells is assumed to form ordered domains more effectively than sphingomyelin due to its greater potential for intermolecular hydrogen bonds and heterogeneous distribution in the sphingomyelin-rich raft-like liquid-ordered phase-segregated membrane domains (Kinoshita et al. 2020). Moreover, reduced sphingosine levels in COVID-19 patients could also increase viral entry since sphingosine prevents binding of SARS-CoV-2 spike to its cellular receptor ACE2 (Edwards et al. 2020).

Our data indicate that pharmacological alterations in membrane composition may serve as a promising target for preventing infection and more severe courses of SARS-CoV-2. Accordingly, via functional inhibition of lysosomal ASM and thus reduction of Cer levels, FIASMAs such as fluoxetine and fluvoxamine have been effective in clinical studies to reduce morbidity and mortality in COVID-19 patients (Hoertel et al. 2021a; Hoertel et al. 2021c). The potential efficacy of these drugs, most of which are approved by the U.S. Food and Drug Administration, most likely minimally toxic and potentially readily available for repurposing, is attributed to their high volume distribution particularly in the lung as the entry site of SARS-CoV-2: the high lung-to-blood concentration gradient of approximately 50 in mice and rats (Cassano et al. 1965) is even excelled in humans (Johnson et al. 2007). Thus, conventional oral administration could be feasible and supported by the expected longer elimination half-life of these drugs due to enrichment in deep compartments such as lysosomes. In addition to reducing infection by prophylactic and early antiviral administration of FIASMAs, these drugs could be also beneficial for later severe courses of COVID-19 due to their expected reduction of the cytokine storm (Lee et al. 2021b) associated with inhibition of ASM activity which is involved in IL-6 (Perry et al. 2014) and TNF-alpha signaling (Schutze et al. 1992).

In addition to targeting ASM, further enzymes involved in the Cer pathway such as NSM, AC or NC could also present potential targets for inhibitory or activating pharmacological interventions. In a preclinical study, administration of anti-Cer antibodies or recombinant NC prevented infection of human nasal epithelial cells with SARS-CoV-2 (Carpinteiro et al. 2020). In contrast, chloroquine, whose early approval for COVID-19 treatment has been revoked due to a lack of benefit in inpatients, leads to an increase in Cer in murine lung cells (Teichgraber et al. 2008) possibly via inhibition of AC typical for amphiphilic lysomotropic agents (Elojeimy et al. 2006) and thus increase in Cer with potentially negative effects. The inhibitory effect of FIASMA-SSRIs on ASM and thus potentially the extent of reduction of Cer levels seems to correlate with the strength of the antiviral effect against SARS-CoV-2 *in vitro* (fluoxetine > paroxetine > fluvoxamine > other SSRIs, (Kornhuber et al. 2021)). Hence, the combination of drugs affecting different enzymes of the Cer pathway to reduce its levels might be even more promising similar to approaches of pharmacological inhibition or depletion of enzymes driving *de novo* Cer synthesis in the area of cardiovascular disease (Choi et al. 2021).

In view of increased ASM and decreased AN/NC activities in COVID-19 patients, it is tempting to speculate that patients with Niemann-Pick disease patients, a rare disorder due to mutations in the ASM encoding gene (Brady 1966), and their heterozygous parents would have a considerably reduced risk of infection with SARS-CoV-2 and less severe course of disease. Contrariwise, patients with Farber disease, another rare sphingolipidosis with defective AC due to mutations in the corresponding gene (Farber 1952), and their heterozygous parents could have an increased risk of COVID-19 with more severe symptoms. However, these hypotheses need to be verified in the future.

In addition to FIASMA action, further sphingolipid related targets for therapeutic approaches have been identified. In a pilot study, 50 healthy volunteers stayed negative for SARS-CoV-2 during 15 days of daily usage of a spray containing alpha-cyclodextrin (depleting sphingolipids and phospholipids from biomembranes) and hydroxytyrosol (anti-viral properties, reducing serum lipids in mice) despite a higher infection risk than the general population. Moreover, two individuals tested positive for SARS-CoV-2 and treated with the spray became negative after half the time compared to four untreated infected even asymptomatic individuals (Ergoren et al. 2020).

Our study is unique with its larger sex-balanced cohort of 200 patients including convalescent patients with antibody levels and novel enzyme activities for NSM and NC. However, it is also subject to several limitations. We have not taken into account the SARS-CoV2-variant causing infection and our data might not be generalizable to all new variants including omicron. Our samples are restricted to one university hospital and to not include healthy or asymptomatic individuals who could be expected to show comparable parameters to convalescent patients. It would be valuable to analyze the sphingolipid pattern during the course of the disease to study the dynamics and to correlate early patterns with progression and outcome to develop predictive markers.

Typical risk factors for a lethal COVID-19 course including age, obesity or hypertension are also associated with the ASM/Cer system (Kornhuber et al. 2021). We were therefore cautious to include age as a cofactor in our statistical models because our groups taken from available hospitalized patients during early pandemic differed significantly in age. Due to a lack of availability, the body mass index as marker of obesity could not be included in our analyses. Future studies should pay attention to collect and integrate full medical data including biochemical laboratory parameters and clinical parameters of disease progression as well as medications which could have additional confounding effects such as FIASMAs.

Due to lacking material, we did not analyze relevant cellular enzymes in peripheral blood mononuclear cells of patients such as lysosomal ASM as well as sphingomyelin synthase (Muhle et al. 2019a) catalyzing the reverse reaction to ASM and thus reducing Cer levels. Peripheral blood cells would also to asses gene expression of a larger panel of sphingolipid metabolizing enzymes, for which biochemical activity assays have not yet been established, as well as splicing patterns. For example, splice variants with dominant-negative effects have been identified for ASM (Rhein et al. 2012) and found to be altered in major depression (Rhein et al. 2017), where mRNA expression also responds to FIASMA treatment (Rhein et al. 2021).

Further biological material such as erythrocyte membranes (Abusukhun et al. 2021), saliva (Frampas et al. 2021) or sebum (Spick et al. 2021), which can be taken easily and painlessly, have been analyzed with respect to metabolomics or lipidomics changes in COVID-19 but not yet for the activity of sphingolipid enzymes which can be determined with less costly and specialized equipment. Such material would be worthy of future consideration for clinical sampling.

Plasma levels of bioactive sphingolipids dihydrosphingosine (sphinganine) and Cer reflected tissue *de novo* sphingolipid synthesis activity as their levels were reduced upon inhibition of serine palmitoyltransferase which catalyzes the first step of Cer *de novo* synthesis by myriocin or upon reduction of substrate free fatty acid levels in the plasma by nicotinic acid. Platelets and erythrocytes also lack *de novo* Cer synthesis, and their levels of sphingolipids also partially followed plasma changes (Blachnio-Zabielska et al. 2016). Thus, these cells could represent additional informative sources of sphingolipid biomarkers with a different time frame of alterations.

Moreover, it would be worthwhile to study the interaction of cellular ceramide levels in lung tissue (thus relying on animal models), peripheral blood cells (as more accessible material for cellular biomarkers) and plasma ceramide levels and their response to FIASMA treatment which primarily targets lysosomal ASM. However, the lysosomal ASM protein is secreted into the circulation upon stress to add up to the constitutively released secretory ASM isoform and is then able to hydrolyze sphingomyelin present at the outer leaflet of membranes (Chung and Claus 2020).

## Conclusions

There is an urgent need for a better understanding of underlying mechanisms, diagnostic markers and highly effective treatments. Our data support the use of FIASMA drugs such as fluoxetine and fluvoxamine, well-tolerated, widely available, orally administered and inexpensive SSRIs in the fight against the COVID-19 pandemic (Hashimoto et al. 2022). They not only reduce Cer levels and thus potentially SARS-CoV-2 entry by functionally inhibiting ASM but -as shown for these and other SSRIs - have also direct effects in controlling inflammation, coagulopathy and the cytokine storm as hallmarks of severe COVID-19 disease (Creeden et al. 2021; Sukhatme et al. 2021). They seem to be effective also against novel variants (Fred et al. 2021).

The combination of sphingolipid and sphingolipid metabolizing enzymes as biomarkers have the potential as a diagnostic marker and may help to prioritize those patients for treatment with medication including approved SARS-CoV-2-neutralizing monoclonal antibodies (Kreuzberger et al. 2021) or newly developed antiviral drugs like molnupiravir (Whitley 2021) or paxlovid (Wang and Yang 2021). These treatments suffer from limited supplies and lower efficacy against variants such as omicron; they are also associated with high cost and side effects and thus would be applied specifically in high-risk cases.

## Data Availability

All data produced in the present study are available upon reasonable request to the authors.

## Authors’ contribution

Conceived and designed the experiments: JK, CM. Provided patient samples: AK, MV, JS, SA. Performed the experiments: CM, FS. Analyzed the data and wrote the paper: CM, BL, JK. Commented on the manuscript and provided intellectual input: all authors.

## Acknowledgements

We thank all patients for providing samples. We are thankful to Alina Bauer for excellent technical support with clinical samples and Daniel Herrmann for excellent technical assistance with the LC-MS/MS analyses.

## Availability of data and materials

Raw and evaluated data are available upon request.

## Compliance with ethical standards

### Ethics approval

The study was approved by the Ethics Committee of the Friedrich-Alexander University Erlangen-Nürnberg (174_20B, April 30, 2020 for patients and 357_19B, October 18^th^, 2019 for convalescent plasma donors).

### Consent to participate

All study participants provided informed consent.

### Consent for publication

The patients provided their written informed consent to use the data for all scientific issues.

