## Supplemental tables and figures for "COVID-19 and its clinical severity are associated with alterations of plasma sphingolipids and enzyme activities of sphingomyelinase and ceramidase"

### Supplementary Material

**Table S1:** Raw data on **a**ctivities of sphingolipid metabolizing enzymes (fmol/h/µl for ASM and NSM, pmol/h/µl for NC) and sphingolipid concentrations (pmol/20µl sample) in COVID-19 patients with mild, moderate and severe symptoms and in convalescent patients recovered from SARS-CoV-2 infection.

| | convalescent | | mild | | moderate | | severe | |
|---|---|---|---|---|---|---|---|---|
| parameter | n | M±SD | n | M±SD | n | M±SD | n | M±SD |
| plasma_ASM | 51 | 84.7 ± 44.7 | 48 | 170.4 ± 105.5 | 50 | 169.3 ± 101.7 | 50 | 156 ± 111.4 |
| plasma_NSM | 51 | 12 ± 8.9 | 48 | 23 ± 19.6 | 50 | 22.3 ± 16.2 | 50 | 73.8 ± 67.5 |
| plasma_NC | 51 | 1.55 ± 0.72 | 48 | 1.24 ± 0.64 | 50 | 1.06 ± 0.56 | 50 | 0.92 ± 0.57 |
| LCB_dHSph | 51 | 1.76 ± 2.3 | 48 | 1.29 ± 1.44 | 49 | 0.81 ± 0.88 | 50 | 0.67 ± 1.02 |
| LCB_Sph | 51 | 6.38 ± 7.19 | 48 | 6.02 ± 6.34 | 49 | 4.19 ± 4.03 | 50 | 4.08 ± 5.9 |
| LCB_S1P | 51 | 23.8 ± 9.7 | 48 | 15.2 ± 8.5 | 49 | 18.1 ± 8.4 | 50 | 15.8 ± 7.1 |
| dHCer_16:0 | 51 | 1.01 ± 0.32 | 48 | 1.13 ± 0.49 | 49 | 1.58 ± 1.84 | 50 | 1.64 ± 0.94 |
| dHCer_18:0 | 51 | 1.49 ± 0.71 | 48 | 2.33 ± 1.56 | 49 | 3.1 ± 2.13 | 50 | 3.01 ± 2.36 |
| dHCer_20:0 | 51 | 2.37 ± 1.09 | 48 | 2.29 ± 1.32 | 49 | 3.08 ± 2.26 | 50 | 2.59 ± 1.93 |
| dHCer_22:0 | 51 | 10.5 ± 3.9 | 48 | 8.7 ± 4.8 | 49 | 9.3 ± 5.8 | 50 | 8.2 ± 5.3 |
| dHCer_24:0 | 51 | 12.8 ± 5.6 | 48 | 9 ± 4.2 | 49 | 8.7 ± 5.8 | 50 | 6.7 ± 3.9 |
| dHCer_24:1 | 51 | 10.8 ± 4.8 | 48 | 11.5 ± 7.5 | 49 | 12.9 ± 8.9 | 50 | 12.4 ± 8.4 |
| dHCer_total | 51 | 39.5 ± 15.2 | 48 | 36 ± 19.7 | 49 | 40.2 ± 26 | 50 | 36.1 ± 22.5 |
| Cer_16:0 | 51 | 11.6 ± 3.9 | 48 | 12.5 ± 5.5 | 49 | 16.8 ± 9.7 | 50 | 21.1 ± 13.3 |
| Cer_18:0 | 51 | 4.74 ± 1.7 | 48 | 7.82 ± 4.18 | 49 | 10.81 ± 6.56 | 50 | 12.87 ± 12.47 |
| Cer_20:0 | 51 | 6.51 ± 2.02 | 48 | 7.06 ± 3.07 | 49 | 9.32 ± 5.15 | 50 | 10.1 ± 8.65 |
| Cer_22:0 | 51 | 43.7 ± 12.8 | 48 | 43.1 ± 16.9 | 49 | 51.8 ± 21.9 | 50 | 52.5 ± 33.1 |
| Cer_24:0 | 51 | 120 ± 34 | 48 | 101 ± 34 | 49 | 108 ± 49 | 50 | 88 ± 43 |
| Cer_24:1 | 51 | 69.3 ± 19.1 | 48 | 83.6 ± 37.9 | 49 | 104.7 ± 48 | 50 | 108.4 ± 63.7 |
| Cer_total | 51 | 257.7 ± 65 | 48 | 257.4 ± 92.3 | 49 | 305 ± 131.9 | 50 | 298.9 ± 165.5 |
| dHSM-16.0 | 51 | 79.3 ± 21.1 | 48 | 67.8 ± 24.6 | 49 | 63.7 ± 25.5 | 50 | 67 ± 32.9 |
| dHSM_18:0 | 51 | 21.6 ± 10.5 | 48 | 30.3 ± 22.2 | 49 | 28.9 ± 19.8 | 50 | 26.4 ± 20.7 |
| dHSM_20:0 | 51 | 17.5 ± 7.8 | 48 | 12.3 ± 6.2 | 49 | 10.6 ± 5.9 | 50 | 9.2 ± 6.9 |
| dHSM_22:0 | 51 | 49.1 ± 23.3 | 48 | 29.3 ± 14.3 | 49 | 24 ± 14 | 50 | 23.8 ± 21.7 |
| dHSM_24:0 | 25 | 10.6 ± 4.7 | 25 | 4.7 ± 1.7 | 25 | 3.9 ± 1.5 | 25 | 3.1 ± 1.8 |
| dHSM_24:1 | 51 | 61.5 ± 29.4 | 48 | 51.4 ± 29.5 | 49 | 42.3 ± 27.3 | 50 | 48.3 ± 43.2 |
| dHSM_total | 51 | 235 ± 85 | 48 | 193 ± 86 | 49 | 171 ± 81 | 50 | 176 ± 111 |
| SM_16:0 | 51 | 1603 ± 178 | 48 | 1530 ± 240 | 49 | 1473 ± 270 | 50 | 1474 ± 282 |
| SM_18:0 | 51 | 370 ± 107 | 48 | 416 ± 198 | 49 | 416 ± 162 | 50 | 327 ± 186 |
| SM_20:0 | 51 | 476 ± 138 | 48 | 350 ± 136 | 49 | 309 ± 122 | 50 | 239 ± 138 |
| SM_22:0 | 51 | 1475 ± 343 | 48 | 1137 ± 327 | 49 | 987 ± 302 | 50 | 909 ± 411 |
| SM_24:0 | 51 | 635 ± 240 | 48 | 411 ± 196 | 49 | 321 ± 150 | 50 | 315 ± 258 |
| SM_24:1 | 51 | 1888 ± 420 | 48 | 1864 ± 496 | 49 | 1716 ± 470 | 50 | 1778 ± 720 |
| SM_total | 51 | 6379 ± 1263 | 48 | 5618 ± 1311 | 49 | 5146 ± 1276 | 50 | 4953 ± 1670 |

ASM acid sphingomyelinase, NC neutral ceramidase, NSM neutral sphingomyelinase, LCB long chain base, dH dehydro, Sph sphingosine, S1P sphingosine-1-phosphate, Cer ceramide, SM sphingomyelin; n sample size (dHSM_24:0 was below detection level in the first set of samples), M±SD mean ± standard deviation

**Table S2:** COVID-19 severity prediction by sphingolipids and sphingolipid metabolism enzymes in combination with sex and age as cofactors (multinomial logistic regression using z-scores of raw values based on the mean and standard deviation of the convalescent group)

| | | | | | convalescent versus | | | | | | | | | mild versus | | | | | | moderate versus | | | p for pairwise comparison | | | | | |
|---|---|---|---|---|---|---|---|---|---|---|---|---|---|---|---|---|---|---|---|---|---|---|---|---|---|---|---|---|
| | | | | | mild | | | moderate | | | severe | | | moderate | | | severe | | | severe | | | conv | conv | conv | mild | mild | mod. |
| | n | Δ | $\chi^2$ | p | OR | LCI | UCI | OR | LCI | UCI | OR | LCI | UCI | OR | LCI | UCI | OR | LCI | UCI | OR | LCI | UCI | mild | mod. | sev | mod. | sev | sev |
| **ASM** | 199 | ↑ | 29.5 | **$1.8 \times 10^{-6}$** | 2.29 | 1.56 | 3.36 | 2.20 | 1.49 | 3.25 | 2.08 | 1.40 | 3.08 | 0.96 | 0.81 | 1.14 | 0.91 | 0.76 | 1.09 | 0.95 | 0.80 | 1.12 | **<0.001** | **<0.001** | **<0.001** | 0.649 | 0.302 | 0.525 |
| **NSM** | 199 | ↑ | 70.0 | **$4.4 \times 10^{-15}$** | 1.64 | 1.17 | 2.32 | 1.58 | 1.11 | 2.24 | 2.42 | 1.68 | 3.48 | 0.96 | 0.78 | 1.18 | 1.47 | 1.21 | 1.79 | 1.53 | 1.26 | 1.87 | **0.005** | **0.011** | **<0.001** | 0.705 | **<0.001** | **<0.001** |
| **NC** | 199 | ↓ | 17.9 | **$4.5 \times 10^{-4}$** | 0.54 | 0.33 | 0.88 | 0.42 | 0.23 | 0.74 | 0.30 | 0.16 | 0.55 | 0.77 | 0.46 | 1.28 | 0.55 | 0.32 | 0.95 | 0.72 | 0.42 | 1.21 | **0.014** | **0.003** | **<0.001** | 0.314 | **0.033** | 0.212 |
| dHSM_16.0 | 198 | ↓ | 12.4 | 0.006 | 0.61 | 0.42 | 0.87 | 0.53 | 0.36 | 0.78 | 0.59 | 0.41 | 0.86 | 0.88 | 0.62 | 1.25 | 0.98 | 0.70 | 1.37 | 1.11 | 0.80 | 1.54 | **0.007** | **0.001** | **0.006** | 0.474 | 0.890 | 0.524 |
| dHSM_18:0 | 198 | | 7.4 | 0.060 | 1.42 | 1.05 | 1.92 | 1.41 | 1.03 | 1.94 | 1.32 | 0.95 | 1.83 | 0.99 | 0.80 | 1.23 | 0.93 | 0.74 | 1.17 | 0.93 | 0.75 | 1.16 | **0.021** | **0.033** | 0.097 | 0.956 | 0.521 | 0.526 |
| **dHSM_20:0** | 198 | ↓ | 25.2 | **$1.4 \times 10^{-5}$** | 0.45 | 0.28 | 0.73 | 0.37 | 0.22 | 0.63 | 0.27 | 0.15 | 0.50 | 0.82 | 0.50 | 1.35 | 0.60 | 0.34 | 1.05 | 0.73 | 0.42 | 1.27 | **0.001** | **<0.001** | **<0.001** | 0.437 | 0.074 | 0.265 |
| **dHSM_22:0** | 198 | ↓ | 27.8 | **$4.0 \times 10^{-6}$** | 0.36 | 0.20 | 0.63 | 0.27 | 0.14 | 0.53 | 0.28 | 0.14 | 0.55 | 0.76 | 0.40 | 1.42 | 0.79 | 0.42 | 1.48 | 1.04 | 0.55 | 1.96 | **<0.001** | **<0.001** | **<0.001** | 0.390 | 0.461 | 0.898 |
| **dHSM_24:0** | 100 | ↓ | 61.2 | **$3.2 \times 10^{-13}$** | 0.01 | 0.00 | 0.14 | 0.00 | 0.00 | 0.07 | 0.00 | 0.00 | 0.02 | 0.37 | 0.08 | 1.77 | 0.09 | 0.01 | 0.57 | 0.23 | 0.04 | 1.40 | **0.001** | **<0.001** | **<0.001** | 0.214 | **0.011** | 0.111 |
| dHSM_24:1 | 198 | | 6.0 | 0.110 | 0.77 | 0.53 | 1.10 | 0.60 | 0.38 | 0.93 | 0.74 | 0.50 | 1.09 | 0.78 | 0.50 | 1.20 | 0.97 | 0.66 | 1.42 | 1.24 | 0.83 | 1.87 | 0.155 | 0.022 | 0.129 | 0.254 | 0.861 | 0.296 |
| dHSM_total | 198 | ↓ | 10.8 | 0.013 | 0.63 | 0.43 | 0.93 | 0.52 | 0.34 | 0.81 | 0.55 | 0.36 | 0.85 | 0.83 | 0.55 | 1.25 | 0.88 | 0.59 | 1.31 | 1.06 | 0.72 | 1.57 | **0.022** | **0.003** | **0.007** | 0.365 | 0.528 | 0.759 |
| SM_16:0 | 198 | ↓ | 14.7 | 0.002 | 0.66 | 0.47 | 0.94 | 0.53 | 0.36 | 0.76 | 0.53 | 0.36 | 0.77 | 0.79 | 0.58 | 1.08 | 0.79 | 0.58 | 1.08 | 1.00 | 0.75 | 1.31 | **0.019** | **0.001** | **0.001** | 0.147 | 0.142 | 0.976 |
| SM_18:0 | 198 | ↓ | 9.2 | 0.027 | 1.13 | 0.86 | 1.48 | 1.14 | 0.85 | 1.54 | 0.78 | 0.55 | 1.10 | 1.02 | 0.79 | 1.31 | 0.69 | 0.51 | 0.94 | 0.68 | 0.51 | 0.91 | 0.393 | 0.378 | 0.151 | 0.904 | **0.017** | **0.009** |
| **SM_20:0** | 198 | ↓ | 57.0 | **$2.6 \times 10^{-12}$** | 0.35 | 0.21 | 0.56 | 0.28 | 0.17 | 0.48 | 0.13 | 0.07 | 0.25 | 0.82 | 0.53 | 1.25 | 0.38 | 0.22 | 0.66 | 0.46 | 0.27 | 0.79 | **<0.001** | **<0.001** | **<0.001** | 0.352 | **0.001** | **0.005** |
| **SM_22:0** | 198 | ↓ | 41.3 | **$5.6 \times 10^{-9}$** | 0.39 | 0.24 | 0.62 | 0.28 | 0.16 | 0.48 | 0.23 | 0.13 | 0.40 | 0.72 | 0.47 | 1.10 | 0.58 | 0.37 | 0.91 | 0.81 | 0.53 | 1.23 | **<0.001** | **<0.001** | **<0.001** | 0.131 | **0.018** | 0.326 |
| **SM_24:0** | 198 | ↓ | 36.2 | **$6.9 \times 10^{-8}$** | 0.40 | 0.25 | 0.65 | 0.26 | 0.14 | 0.47 | 0.26 | 0.14 | 0.48 | 0.65 | 0.38 | 1.10 | 0.66 | 0.38 | 1.12 | 1.01 | 0.58 | 1.77 | **<0.001** | **<0.001** | **<0.001** | 0.107 | 0.120 | 0.962 |
| SM_24:1 | 198 | | 5.5 | 0.136 | 0.87 | 0.63 | 1.22 | 0.66 | 0.45 | 0.96 | 0.72 | 0.50 | 1.05 | 0.75 | 0.54 | 1.06 | 0.83 | 0.59 | 1.15 | 1.10 | 0.80 | 1.50 | 0.430 | **0.030** | 0.090 | 0.101 | 0.266 | 0.557 |
| **SM_total** | 198 | ↓ | 24.9 | **$1.6 \times 10^{-5}$** | 0.54 | 0.36 | 0.81 | 0.39 | 0.25 | 0.62 | 0.35 | 0.22 | 0.56 | 0.73 | 0.49 | 1.08 | 0.65 | 0.43 | 0.97 | 0.88 | 0.61 | 1.29 | **0.003** | **<0.001** | **<0.001** | 0.115 | **0.034** | 0.522 |
| **dHCer_16:0** | 198 | ↑ | 17.9 | **$4.6 \times 10^{-4}$** | 1.17 | 0.87 | 1.57 | 1.58 | 1.18 | 2.11 | 1.60 | 1.20 | 2.14 | 1.35 | 1.06 | 1.72 | 1.37 | 1.08 | 1.75 | 1.02 | 0.93 | 1.11 | 0.308 | **0.002** | **0.002** | **0.014** | **0.010** | 0.729 |
| **dHCer_18:0** | 198 | ↑ | 26.9 | **$6.1 \times 10^{-6}$** | 1.72 | 1.21 | 2.46 | 2.02 | 1.40 | 2.90 | 1.99 | 1.38 | 2.87 | 1.17 | 0.99 | 1.39 | 1.16 | 0.98 | 1.37 | 0.99 | 0.87 | 1.12 | **0.003** | **<0.001** | **<0.001** | 0.061 | 0.091 | 0.843 |
| dHCer_20:0 | 198 | | 5.7 | 0.130 | 1.02 | 0.72 | 1.45 | 1.36 | 0.97 | 1.91 | 1.19 | 0.83 | 1.69 | 1.33 | 0.99 | 1.80 | 1.16 | 0.85 | 1.59 | 0.87 | 0.70 | 1.09 | 0.901 | 0.072 | 0.341 | 0.059 | 0.347 | 0.228 |
| dHCer_22:0 | 198 | | 3.7 | 0.295 | 0.75 | 0.53 | 1.07 | 0.87 | 0.62 | 1.22 | 0.74 | 0.51 | 1.07 | 1.15 | 0.82 | 1.60 | 0.98 | 0.69 | 1.40 | 0.85 | 0.63 | 1.16 | 0.114 | 0.405 | 0.108 | 0.419 | 0.915 | 0.319 |
| **dHCer_24:0** | 198 | ↓ | 22.1 | **$6.2 \times 10^{-5}$** | 0.47 | 0.28 | 0.79 | 0.50 | 0.29 | 0.84 | 0.26 | 0.14 | 0.50 | 1.06 | 0.64 | 1.74 | 0.56 | 0.31 | 1.02 | 0.53 | 0.30 | 0.94 | **0.004** | **0.009** | **<0.001** | 0.826 | 0.058 | **0.029** |
| dHCer_24:1 | 198 | | 1.0 | 0.792 | 1.07 | 0.78 | 1.47 | 1.17 | 0.85 | 1.60 | 1.12 | 0.81 | 1.55 | 1.09 | 0.84 | 1.41 | 1.05 | 0.81 | 1.37 | 0.96 | 0.77 | 1.20 | 0.684 | 0.344 | 0.486 | 0.510 | 0.715 | 0.738 |
| dHCer_total | 198 | | 1.4 | 0.707 | 0.89 | 0.64 | 1.23 | 1.03 | 0.75 | 1.41 | 0.91 | 0.65 | 1.28 | 1.16 | 0.86 | 1.57 | 1.03 | 0.74 | 1.42 | 0.89 | 0.68 | 1.16 | 0.477 | 0.853 | 0.594 | 0.342 | 0.872 | 0.383 |

| | | | | | | | | | | | | | | | | | | | | | | | | | | | | |
|---|---|---|---|---|---|---|---|---|---|---|---|---|---|---|---|---|---|---|---|---|---|---|---|---|---|---|---|---|
| **Cer_16:0** | 198 | ↑ | 27.0 | **$5.8 \times 10^{-6}$** | 1.12 | 0.76 | 1.64 | 1.71 | 1.18 | 2.48 | 1.96 | 1.35 | 2.85 | 1.53 | 1.13 | 2.09 | 1.76 | 1.29 | 2.40 | 1.15 | 0.98 | 1.34 | 0.571 | **0.004** | **<0.001** | **0.007** | **<0.001** | 0.081 |
| **Cer_18:0** | 198 | ↑ | 49.8 | **$8.7 \times 10^{-11}$** | 2.00 | 1.41 | 2.82 | 2.41 | 1.69 | 3.43 | 2.51 | 1.76 | 3.57 | 1.20 | 1.02 | 1.42 | 1.25 | 1.06 | 1.48 | 1.04 | 0.97 | 1.12 | **<0.001** | **<0.001** | **<0.001** | **0.026** | **0.007** | 0.291 |
| Cer_20:0 | 198 | ↑ | 11.8 | 0.008 | 1.12 | 0.82 | 1.53 | 1.43 | 1.05 | 1.94 | 1.48 | 1.09 | 2.02 | 1.28 | 1.00 | 1.63 | 1.32 | 1.03 | 1.69 | 1.04 | 0.92 | 1.16 | 0.476 | **0.022** | **0.012** | 0.052 | **0.026** | 0.545 |
| Cer_22:0 | 198 | ↑ | 4.7 | 0.194 | 0.94 | 0.68 | 1.29 | 1.19 | 0.89 | 1.59 | 1.22 | 0.91 | 1.64 | 1.27 | 0.95 | 1.68 | 1.30 | 0.98 | 1.72 | 1.02 | 0.85 | 1.24 | 0.698 | 0.251 | 0.192 | 0.102 | 0.071 | 0.801 |
| **Cer_24:0** | 198 | ↓ | 16.5 | **$8.9 \times 10^{-4}$** | 0.59 | 0.40 | 0.88 | 0.69 | 0.47 | 1.01 | 0.44 | 0.29 | 0.69 | 1.16 | 0.81 | 1.67 | 0.75 | 0.50 | 1.12 | 0.65 | 0.45 | 0.94 | **0.009** | 0.054 | **<0.001** | 0.423 | 0.163 | **0.021** |
| Cer_24:1 | 198 | ↑ | 11.0 | 0.012 | 1.27 | 0.94 | 1.71 | 1.49 | 1.10 | 2.02 | 1.52 | 1.12 | 2.07 | 1.18 | 0.95 | 1.45 | 1.20 | 0.98 | 1.48 | 1.02 | 0.89 | 1.17 | 0.121 | **0.010** | **0.007** | 0.129 | 0.084 | 0.748 |
| Cer_total | 198 | | 2.8 | 0.416 | 0.92 | 0.69 | 1.24 | 1.12 | 0.86 | 1.46 | 1.11 | 0.85 | 1.45 | 1.21 | 0.94 | 1.57 | 1.20 | 0.92 | 1.55 | 0.99 | 0.82 | 1.19 | 0.598 | 0.386 | 0.454 | 0.142 | 0.177 | 0.882 |
| LCB_dHSph | 198 | ↓ | 12.9 | 0.005 | 0.77 | 0.45 | 1.32 | 0.36 | 0.16 | 0.81 | 0.25 | 0.10 | 0.66 | 0.47 | 0.22 | 1.03 | 0.33 | 0.13 | 0.83 | 0.70 | 0.26 | 1.86 | 0.344 | **0.013** | **0.005** | 0.059 | **0.019** | 0.474 |
| LCB_Sph | 198 | ↓ | 6.6 | 0.087 | 0.93 | 0.60 | 1.44 | 0.57 | 0.32 | 1.02 | 0.55 | 0.31 | 1.00 | 0.62 | 0.36 | 1.06 | 0.59 | 0.34 | 1.04 | 0.97 | 0.53 | 1.77 | 0.751 | 0.058 | 0.051 | 0.078 | 0.067 | 0.909 |
| **LCB_S1P** | 198 | ↓ | 17.7 | **$5.1 \times 10^{-4}$** | 0.37 | 0.22 | 0.62 | 0.65 | 0.40 | 1.06 | 0.48 | 0.28 | 0.82 | 1.77 | 1.05 | 2.99 | 1.30 | 0.75 | 2.25 | 0.74 | 0.46 | 1.18 | **<0.001** | 0.082 | **0.007** | **0.033** | 0.342 | 0.206 |

ASM acid sphingomyelinase, NC neutral ceramidase, NSM neutral sphingomyelinase, LCB long chain base, dH dehydro, Sph sphingosine, S1P sphingosine-1-phosphate, Cer ceramide, SM sphingomyelin; n total sample size; for $p<0.05$: ↑ increased and ↓ decreased in severely affected patients versus convalescent patients, parameter and p value in bold for those remaining significant after Bonferroni correction for multiple testing; OR odds ratio; LCI lower and UCI upper confidence intervals; $p<0.05$ in bold for pair-wise comparison;

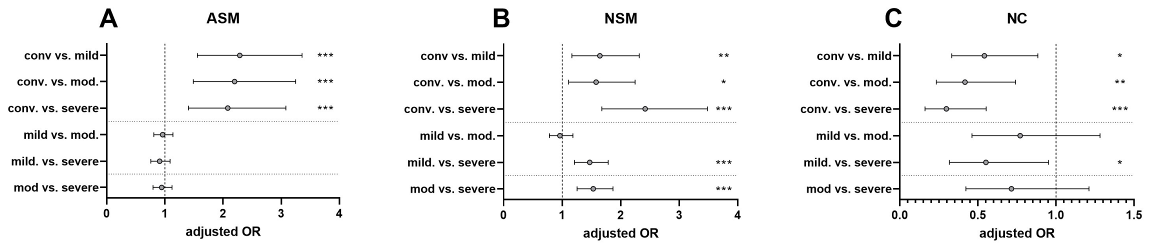


***Figure S3****: Odds ratios and significance of pair-wise comparisons of patients with mild, moderate and severe symptoms with convalescent subjects (odds ratios with 95% confidence intervals as error bars) for sphingolipid metabolism enzymes from multinomial regression analysis of z-scores (calculated from raw values based on mean and standard deviation of the convalescent group) taking into account sex and age (see Table S2 for statistical information). *p < 0.05, **p < 0.01, ***p < 0.001*

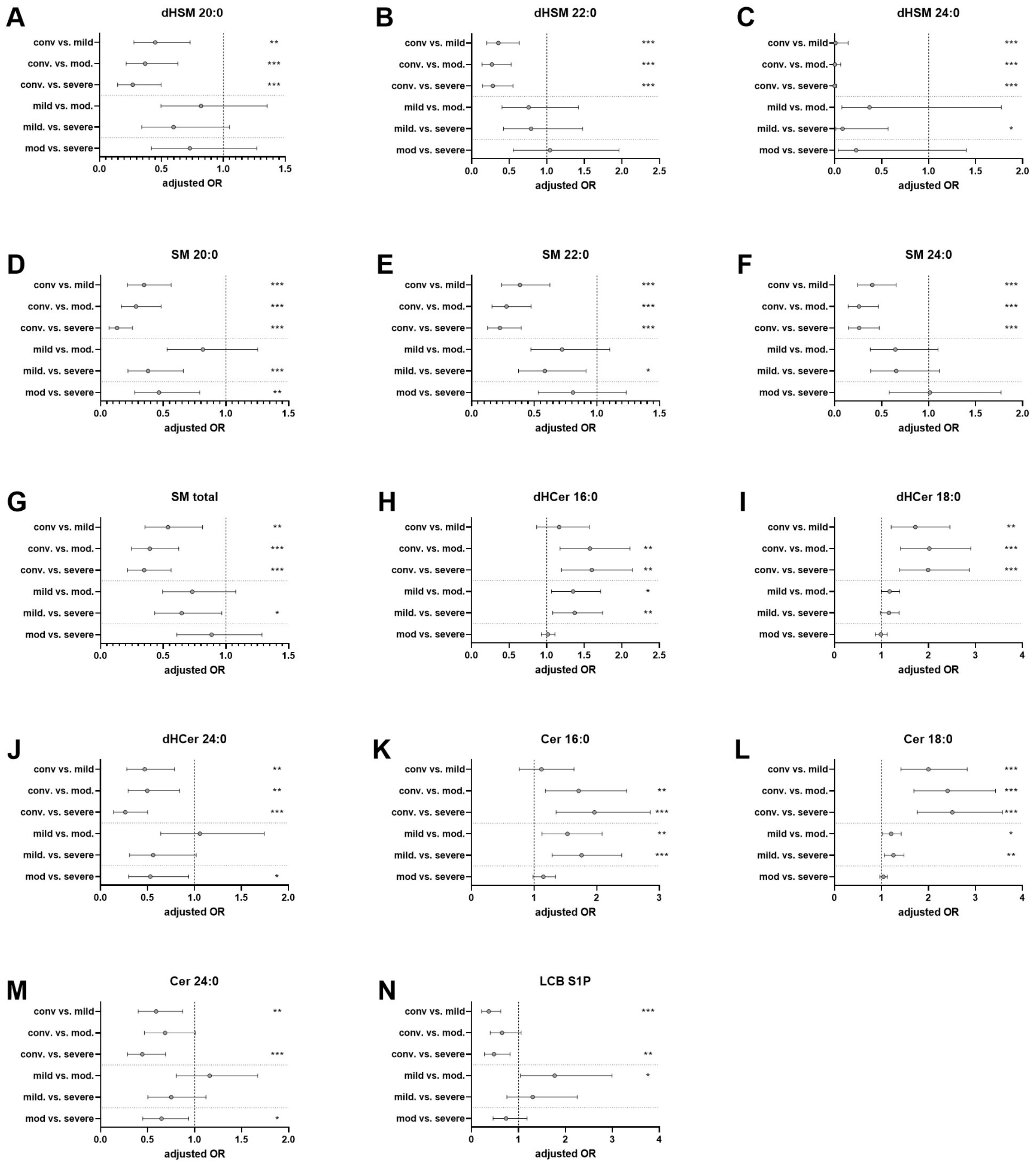


***Figure S4**: Odds ratios and significance of pair-wise comparisons of patients with mild, moderate and severe symptoms with convalescent subjects (odds ratios with 95% confidence intervals as error bars) for sphingolipids (LC-MS/MS data, see Figure 1) from multinomial regression analysis of z-scores (calculated from raw values based on mean and standard deviation of the convalescent group) taking into account sex and age (see Table S2 for statistical information). *p < 0.05, **p < 0.01, ***p < 0.001*

**Table S5**: Sex-separated COVID-19 severity prediction by sphingolipids and sphingolipid metabolism enzymes in combination with age as predictor (linear regression model coding 0 = convalecent, 1 = mild, 2 = moderate, 3 = severe symptoms).

| | **male patients** | | | | | **female patients** | | | | |
|---|---|---|---|---|---|---|---|---|---|---|
| | **n** | **change** | **standar-dized B** | **p for change in F** | **change in $R^2$** | **n** | **change** | **standar-dized B** | **p for change in F** | **change in $R^2$** |
| plasma_ASM | 125 | ns | 0.109 | 0.171 | 0.011 | 74 | ↑ | 0.185 | 0.042 | 0.033 |
| plasma_NC | 125 | ↓ | -0.172 | 0.026 | 0.029 | 74 | ↓ | -0.325 | **6.83 × $10^{-4}$** | 0.087 |
| plasma_NSM | 125 | ↑ | 0.403 | **1.08 × $10^{-7}$** | 0.153 | 74 | ↑ | 0.365 | **2.79 × $10^{-5}$** | 0.127 |
| LCB_dhSph | 125 | ↓ | -0.260 | **6.64 × $10^{-4}$** | 0.067 | 73 | ns | -0.025 | 0.793 | 0.001 |
| LCB_Sph | 125 | ↓ | -0.201 | 0.009 | 0.041 | 73 | ns | 0.036 | 0.701 | 0.001 |
| LCB_S1P | 125 | ns | -0.145 | 0.069 | 0.020 | 73 | ns | -0.125 | 0.181 | 0.015 |
| dhCer_16:0 | 125 | ↑ | 0.186 | 0.016 | 0.034 | 73 | ↑ | 0.257 | 0.005 | 0.063 |
| dhCer_18:0 | 125 | ↑ | 0.257 | **8.32 × $10^{-4}$** | 0.065 | 73 | ns | 0.161 | 0.085 | 0.024 |
| dhCer_20:0 | 125 | | 0.101 | 0.192 | 0.010 | 73 | | 0.002 | 0.983 | 0.000 |
| dhCer_22:0 | 125 | | -0.073 | 0.353 | 0.005 | 73 | | -0.076 | 0.407 | 0.006 |
| dhCer_24:0 | 125 | ↓ | -0.276 | **4.95 × $10^{-4}$** | 0.070 | 73 | ↓ | -0.215 | 0.022 | 0.042 |
| dhCer_24:1 | 125 | | 0.059 | 0.447 | 0.004 | 73 | | 0.017 | 0.852 | 0.000 |
| dhCer_total | 125 | | -0.005 | 0.947 | 0.000 | 73 | | -0.032 | 0.727 | 0.001 |
| Cer_16:0 | 125 | ↑ | 0.271 | **4.49 × $10^{-4}$** | 0.071 | 73 | ↑ | 0.330 | **3.62 × $10^{-4}$** | 0.096 |
| Cer_18:0 | 125 | ↑ | 0.299 | **9.37 × $10^{-5}$** | 0.087 | 73 | ↑ | 0.278 | 0.004 | 0.066 |
| Cer_20:0 | 125 | ↑ | 0.216 | 0.005 | 0.046 | 73 | ns | 0.095 | 0.318 | 0.008 |
| Cer_22:0 | 125 | ↑ | 0.152 | 0.049 | 0.023 | 73 | ns | 0.028 | 0.767 | 0.001 |
| Cer_24:0 | 125 | ↓ | -0.182 | 0.019 | 0.033 | 73 | ↓ | -0.210 | 0.019 | 0.043 |
| Cer_24:1 | 125 | ↑ | 0.208 | 0.008 | 0.041 | 73 | ns | 0.130 | 0.194 | 0.014 |
| Cer_total | 125 | | 0.104 | 0.182 | 0.011 | 73 | | 0.021 | 0.827 | 0.000 |
| dhSM_16.0 | 125 | ns | -0.134 | 0.084 | 0.018 | 73 | ns | -0.133 | 0.142 | 0.018 |
| dhSM_18:0 | 125 | | 0.105 | 0.178 | 0.011 | 73 | | 0.012 | 0.892 | 0.000 |
| dhSM_20:0 | 125 | ↓ | -0.298 | **1.19 × $10^{-4}$** | 0.085 | 73 | ↓ | -0.239 | 0.011 | 0.051 |
| dhSM_22:0 | 125 | ↓ | -0.270 | **6.42 × $10^{-4}$** | 0.068 | 73 | ↓ | -0.271 | 0.006 | 0.059 |
| dhSM_24:0 | 58 | ↓ | -0.708 | **4.64 × $10^{-10}$** | 0.425 | 42 | ns | -0.265 | 0.073 | 0.050 |
| dhSM_24:1 | 125 | | -0.058 | 0.454 | 0.003 | 73 | | -0.139 | 0.136 | 0.018 |
| dhSM_total | 125 | ns | -0.147 | 0.059 | 0.021 | 73 | ns | -0.155 | 0.093 | 0.023 |
| SM_16:0 | 125 | ↓ | -0.169 | 0.029 | 0.028 | 73 | ↓ | -0.195 | 0.030 | 0.038 |
| SM_18:0 | 125 | | -0.051 | 0.512 | 0.003 | 73 | | -0.108 | 0.234 | 0.012 |
| SM_20:0 | 125 | ↓ | -0.436 | **5.63 × $10^{-9}$** | 0.180 | 73 | ↓ | -0.364 | **1.08 × $10^{-4}$** | 0.112 |
| SM_22:0 | 125 | ↓ | -0.326 | **3.80 × $10^{-5}$** | 0.096 | 73 | ↓ | -0.404 | **3.41 × $10^{-5}$** | 0.126 |
| SM_24:0 | 125 | ↓ | -0.314 | **6.02 × $10^{-5}$** | 0.092 | 73 | ↓ | -0.349 | **4.56 × $10^{-4}$** | 0.093 |
| SM_24:1 | 125 | | -0.025 | 0.745 | 0.001 | 73 | ↓ | -0.209 | 0.021 | 0.043 |
| SM_total | 125 | ↓ | -0.231 | 0.003 | 0.052 | 73 | ↓ | -0.302 | **1.05 × $10^{-3}$** | 0.082 |

ASM acid sphingomyelinase, NC neutral ceramidase, NSM neutral sphingomyelinase, LCB long chain base, dH dihydro, Sph sphingosine, S1P sphingosine-1-phosphate, Cer ceramide, SM sphingomyelin; n total sample size; for $p < 0.05$: ↑ increased and ↓ decreased in severely affected patients versus convalescent patients, ; all these remained $p < 0.05$ except for dHCer 16:0 for males in Bootstrap analysis; ns not nominally significant although it was significant in the entire cohort; p value in bold for those remaining significant after Bonferroni correction for multiple hypothesis testing.
